# Coronavirus vaccination rates in cultural areas in Germany

**DOI:** 10.1101/2021.07.28.21261246

**Authors:** Claudia Spahn, Anna Maria Hipp, Bernhard Richter, Manfred Nusseck

**Affiliations:** Freiburg Institute for Musicians’ Medicine, University Medical Center Freiburg, University of Music Freiburg, Medical Faculty of the Albert Ludwigs University Freiburg, Freiburg Center for Research and Teaching in Music, Germany

**Author notes:** **Correspondence:** Prof. Dr. Dr. Claudia Spahn.

## Abstract

Vaccination rates can provide useful information about potential risks of infection in a given population. In this study, the vaccination rates and attitudes toward vaccination in cultural sectors, i.e. music areas, have been investigated. In total, 4341 persons in four different areas including visitors of classical music and musicals as well as professional and amateur musicians participated in this survey. Results showed rates of 86% recovered or vaccinated at least once and 54.5% fully vaccinated. These vaccination rates were considerably higher compared to the general population. Vaccination hesitancy was half that of the general population at 6.4%. The findings of this large sample indicate that in the field of music culture there is a high vaccination rate and a low rejection of vaccination on the part of the audience and the performers. The risk of infection can therefore be assumed as very low and the opening of cultural events can thus be recommended.

## Introduction

The impact of COVID-19 had a tremendous effect on cultural areas. Music concerts, theatres, festivals and nearly all other kinds of live performances in front of an audience were governmentally restricted in accordance with the present pandemic situation. The same holds true for rehearsals with large groups of musicians, i.e. for singers and brass instrumentalists. Under the circumstances of dropped incidence rates, some cultural areas have reopened with limited access. In order to be able to carry out the possibility of further openings, the infection situation as well as the level of vaccination in the people involved in these events should be investigated. Therefore, the focus of this study was on vaccination rates among the population in specific cultural areas.

Current vaccines against COVID-19 have been shown to be highly effective (Baden et al., 2021; Haas et al., 2021; Lopez Bernal et al., 2021; Polack et al., 2020). These studies have shown that vaccination dramatically reduces the spread of the virus. It is therefore important to reach a high degree of vaccinated people to reduce the incidence of infection (Evans & Jewell, 2021). Above a certain threshold of the proportion vaccinated in the population, general immunity can be achieved, known as herd immunity (Fine et al., 2011). Models expect to reach herd immunity if at least 66% of the population is vaccinated (MacIntyre et al., 2021). Such high rates of vaccination are currently found in certain occupational groups, such as healthcare workers with rates between 72.9% (Azamgarhi et al., 2021) and 89% (Hall et al., 2021).

However, the vaccination rates in the general population are normally considerably lower. At the time of the present study (June/July 2021), the epidemiologic situation of vaccination rates in Germany was approximately 55% of the population who had been vaccinated at least once and approximately 37% who were fully vaccinated (RKI July 2021).

On the other hand, the effectiveness of vaccination depends on people’s willingness to be vaccinated. Vaccine hesitancy can limit the benefits of vaccination on stopping the spread of the virus. The percentage of hesitaters varied by country (Neumann-Böhme et al., 2020). In Germany, a recent survey found that the percentages were at 6.1% of undecided and at 6.3% of opposed people (RKI June 2021).

In cultural events, it was expected to find vaccination rates similar to the general population.

For participation in cultural events, it is necessary to be either completely immunized through a certified recovery from a COVID-19 infection or through a full vaccination (the second shot must be at least two weeks prior) or to report a negative test (taken within less than 24h).

Therefore, it seems highly important to know the vaccination rates in cultural areas.

The goal of this study was to evaluate the vaccination rates in cultural areas. For this, a specification in the music area was chosen. A distinction was made between visitors of music events and active musicians. In addition, a further subdivision was made into professional and amateur areas. This resulted in four focus areas of either attendees at professional and amateur live music events or professional and amateur musicians. The participants were asked to indicate their vaccination status. The results were compared with general vaccination rates and provide insights into the risk of infection at cultural music events.

## Materials and methods

### Participants

A total of N = 4341 persons took part in this survey (Figure 1). The sample consists of 58.1% female, 41.7% male and 0.2% divers participated. The group of visitors showed a higher female distribution (in classical music events 69.8% and in amateur music events 60.5%). The group of musicians were rather equally distributed between female and male musicians (female professional musicians: 47.0%; female amateur musicians: 47.4%).

**Figure 1:**
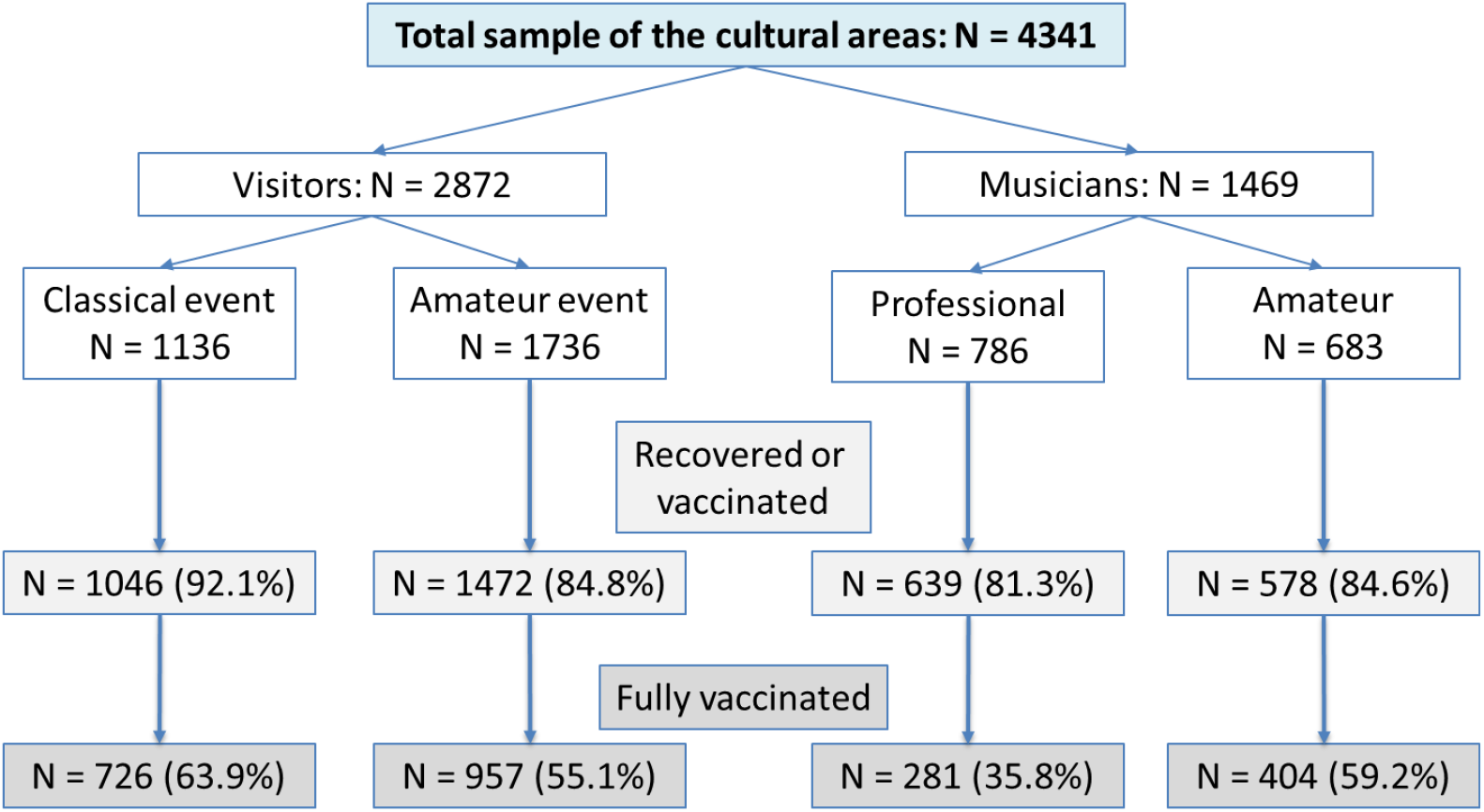
Vaccination rates of the different samples in the cultural areas

In the distribution of the age groups, the largest sizes were found in the groups 18 to 26 years (21%), 46 to 55 years (18%) and 56 to 65 years (20%). The smallest age group was the group of >80 years (3.6%). The other age groups had similar sizes of about 10%. There was a significant distribution difference in the age groups between the focus areas (χ^2^ = 1726.3; p < 0.001) with older participants in the group of visitors of classical music events and the younger participants in the group of the professional musicians.

### Questionnaire

The questionnaire consisted of general questions about age (through age groups) and gender and specific questions about the vaccination status. First, it asked if the person has had a COVID-19 infection (yes/no). If the next question of being vaccinated (yes/no) was positively answered, the participants had to state if they were fully vaccinated, i.e. two shots (or one with J&J). If they were not vaccinated, they were asked if they already made a vaccination date. If not, they were asked to state if they had a vaccination readiness or if they were either undecided or dismissive in being vaccinated. The latter two were also asked about their reasons for not wanting to be vaccinated.

### Study procedure

The study was conducted as an online survey and with print versions of the questionnaire. Participants could decide which version they prefer. On the title page of the print version and on the first page in the online survey, participants were informed that the questionnaire was anonymous and that participation was voluntary. By filling in the questionnaire, they agreed for participation and the scientific use of their data. The study was ethically approved by the Ethics Committee of the University Medical Center Freiburg.

At the music events, the questionnaire was distributed to the visitors with the request to fill it out. The classical music events were operas and ballets. The amateur music events were musicals performed by amateur performers. The events were presented in renowned venues. The audience bought tickets for these events.

The musicians were music students and music teachers at the University of Music Freiburg for the professional sector and brass ensembles and orchestras of the amateur sector. The amateur musicians were asked in rehearsals to participate in the study. In all situations, the applicable hygiene regulations were observed.

### Statistics

The statistical analyses were performed using SPSS version 27 (IBM, Chicago, IL). The data were summarized by using descriptive statistics. Categorical data were compared using cross tables and Pearson’s χ^2^ tests. The level of significance was set at p = 0.05.

## Results

Of the total sample, 86% reported being recovered or at least vaccinated once and 54.5% were fully vaccinated (Figure 1). There were significant differences in the vaccination rates between the cultural areas (χ^2^ = 180.1, p < 0.001). The highest vaccination rates were found in the group of visitors of classical events with 92.1% recovered or at least vaccinated once and with 63.9% fully vaccinated. The lowest rates were reported in the group of professional musicians (81.3% and 35.8% respectively).

The distribution of the vaccination rates within each age group are shown in Figure 2. The number of participants who were recovered or at least vaccinated once was lowest for the youngest age group (18-26 years; 71.4%). All other age groups were above 80%. The number of fully vaccinated participants increased steadily across the age groups from 31.8% in the age group 18-26 years to the highest amount in the age group >80 years with 91.5%.

**Figure 2:**
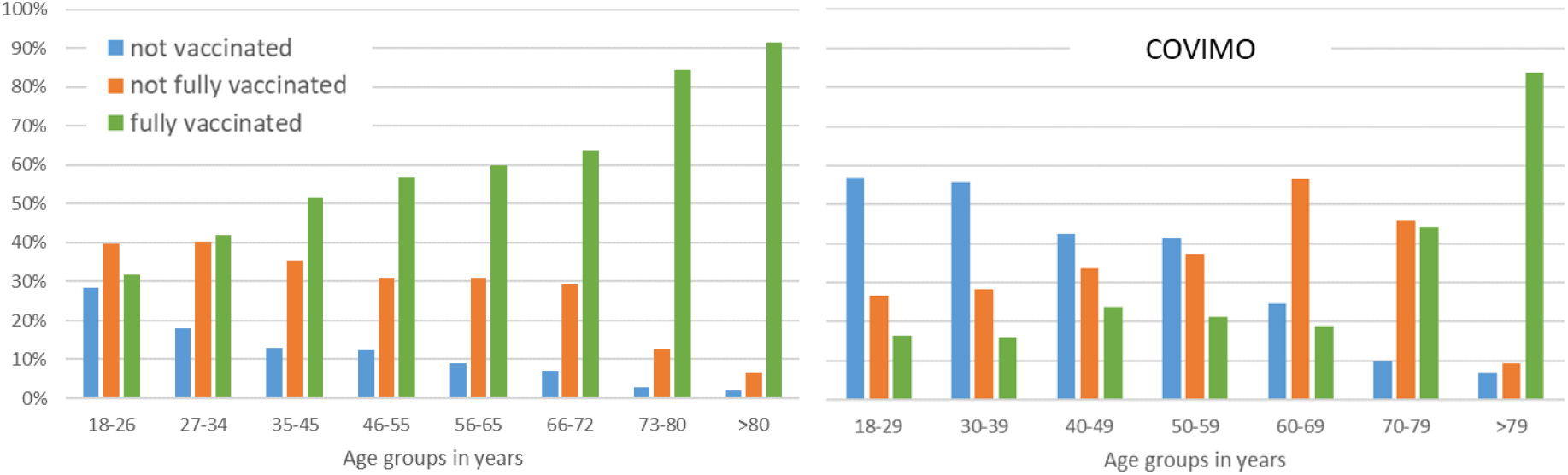
Vaccination rates by age groups (left: findings in this study, N = 4341; right: general population in Germany provided by the RKI, June 2021, N = 3004)

In comparison, the vaccination rates of the general population (RKI, June 2021) are presented to the right of the rates of this study. In every age group, the number of fully vaccinated participants in this study was higher than in the general population and except for the participants older than 79 years, the rates were approximately twice as high. The number of unvaccinated participants in this study was below 30% in the age group 18-26 and even lower than 20% for all other age groups. In the general population, the rate of unvaccinated persons was lower than 30% only in the age groups over 60 years. The rate was more than twice as high as the rate in this study for all age groups.

The overall proportion of participants in this study who showed vaccination hesitancy was 6.4%. This amount can be divided into participants who were undecided about getting vaccinated (N = 143; 3.3%) and participants who decline vaccination (N = 133, 3.1%). Both rates were significantly lower compared to the general population (RKI, June 2021) of undecided persons (6.1%; χ^2^ = 32.8, p < 0.001) and of vaccination deniers (6.3%; χ^2^ = 43.1, p < 0.001). The reasons for vaccination hesitancy were mainly the fear of side effects, the uncertainty of the vaccines in the long-term perspective or that they simply refuse any kind of vaccination.

## Discussion

In this study, the vaccination rates of the population in different music areas were evaluated. The results showed very high vaccination rates in all areas of music culture. In comparison with the general population, the rates in the cultural areas were considerably higher. The rates were to the rates of healthcare workers (Azamgarhi et al., 2021; Hall et al., 2021) and teachers (RKI June 2021).

The vaccination rates in the cultural areas exceeded the rates of the general population in all age groups. In Germany, there was no compulsory vaccination and specific people were prioritized for vaccination, including the older population. As the attendees of the cultural events are on average above mid-age, the old age of the visitors could explain the high vaccination rates. However, the results showed that even in younger age groups the vaccination rates were extraordinarily high.

Especially vaccination hesitancy is very low compared to the general population. This indicates a high willingness for vaccination in the population of cultural areas that is very important in times of free access to vaccinations. The findings also indicate that the population in cultural areas is mainly composed of people who care for their safety and attend music events only with a certain degree of immunization.

In conclusion, the available data on a large sample of audiences in the classical and musical fields, as well as among active musicians in the professional and amateur fields, show that a high vaccination rate and willingness to be vaccinated can be expected in the field of musical culture.

According to the results of the study, it can be expected that more than 90% of the participants in the cultural sector will be fully vaccinated in the fall. This provides favorable conditions for continuing to keep the music culture field open in the fall of 2021, even under potentially increasing incidence rates.

## Data Availability

The data can be requested by the corresponding author

## Author Contributions

All authors considerably participated in the conceptualization and execution of the study. They also contributed to interpreting the results and revising the draft. The corresponding author attests that all listed authors meet authorship criteria.

## Institutional Review Board Statement

This study has been supervised for the ethical guidelines by the Ethics Committee of the University Medical Center Freiburg.

## Data Availability Statement

The data can be requested by the corresponding author.

## Literature

Azamgarhi, T., Hodgkinson, M., Shah, A., Skinner, J. A., Hauptmannova, I., Briggs, T. W. R., & Warren, S. (2021). BNT162b2 vaccine uptake and effectiveness in UK healthcare workers – a single centre cohort study. Nature Communications, 12(1), 3698. https://doi.org/10.1038/s41467-021-23927-x

Baden, L. R., El Sahly, H. M., Essink, B., Kotloff, K., Frey, S., Novak, R., Diemert, D., Spector, S. A., Rouphael, N., Creech, C. B., McGettigan, J., Khetan, S., Segall, N., Solis, J., Brosz, A., Fierro, C., Schwartz, H., Neuzil, K., Corey, L., … Zaks, T. (2021). Efficacy and Safety of the mRNA-1273 SARS-CoV-2 Vaccine. New England Journal of Medicine, 384(5), 403–416. https://doi.org/10.1056/NEJMoa2035389

Evans, S. J. W., & Jewell, N. P. (2021). Vaccine Effectiveness Studies in the Field. New England Journal of Medicine, NEJMe2110605. https://doi.org/10.1056/NEJMe2110605

Fine, P., Eames, K., & Heymann, D. L. (2011). “Herd Immunity”: A Rough Guide. Clinical Infectious Diseases, 52(7), 911–916. https://doi.org/10.1093/cid/cir007

Haas, E. J., Angulo, F. J., McLaughlin, J. M., Anis, E., Singer, S. R., Khan, F., Brooks, N., Smaja, M., Mircus, G., Pan, K., Southern, J., Swerdlow, D. L., Jodar, L., Levy, Y., & Alroy-Preis, S. (2021). Impact and effectiveness of mRNA BNT162b2 vaccine against SARS-CoV-2 infections and COVID-19 cases, hospitalisations, and deaths following a nationwide vaccination campaign in Israel: An observational study using national surveillance data. The Lancet, 397(10287), 1819–1829. https://doi.org/10.1016/S0140-6736(21)00947-8

Hall, V. J., Foulkes, S., Saei, A., Andrews, N., Oguti, B., Charlett, A., Wellington, E., Stowe, J., Gillson, N., Atti, A., Islam, J., Karagiannis, I., Munro, K., Khawam, J., Group, T. S. S., Chand, M. A., Brown, C., Ramsay, M. E., Bernal, J. L., & Hopkins, S. (2021). Effectiveness of BNT162b2 mRNA Vaccine Against Infection and COVID-19 Vaccine Coverage in Healthcare Workers in England, Multicentre Prospective Cohort Study (the SIREN Study). SSRN Electronic Journal. https://doi.org/10.2139/ssrn.3790399

Lopez Bernal, J., Andrews, N., Gower, C., Gallagher, E., Simmons, R., Thelwall, S., Stowe, J., Tessier, E., Groves, N., Dabrera, G., Myers, R., Campbell, C. N. J., Amirthalingam, G., Edmunds, M., Zambon, M., Brown, K. E., Hopkins, S., Chand, M., & Ramsay, M. (2021). Effectiveness of Covid-19 Vaccines against the B.1.617.2 (Delta) Variant. New England Journal of Medicine, NEJMoa2108891. https://doi.org/10.1056/NEJMoa2108891

MacIntyre, C. R., Costantino, V., & Trent, M. (2021). Modelling of COVID-19 vaccination strategies and herd immunity, in scenarios of limited and full vaccine supply in NSW, Australia. Vaccine, S0264410X21005016. https://doi.org/10.1016/j.vaccine.2021.04.042

Neumann-Böhme, S., Varghese, N. E., Sabat, I., Barros, P. P., Brouwer, W., van Exel, J., Schreyögg, J., & Stargardt, T. (2020). Once we have it, will we use it? A European survey on willingness to be vaccinated against COVID-19. The European Journal of Health Economics, 21(7), 977–982. https://doi.org/10.1007/s10198-020-01208-6

Polack, F. P., Thomas, S. J., Kitchin, N., Absalon, J., Gurtman, A., Lockhart, S., Perez, J. L., Pérez Marc, G., Moreira, E. D., Zerbini, C., Bailey, R., Swanson, K. A., Roychoudhury, S., Koury, K., Li, P., Kalina, W. V., Cooper, D., Frenck, R. W., Hammitt, L. L., … Gruber, W. C. (2020). Safety and Efficacy of the BNT162b2 mRNA Covid-19 Vaccine. New England Journal of Medicine, 383(27), 2603–2615. https://doi.org/10.1056/NEJMoa2034577

RKI (June 2021) COVID-19 Impfquoten-Monitoring in Deutschland (COVIMO). 5th survey (May-June 2021). https://www.rki.de/DE/Content/InfAZ/N/Neuartiges_Coronavirus/Projekte_RKI/covimo_studie_Ergebni sse.html

RKI (July 2021) Coronavirus Disease 2019 (COVID-19) - Situation Report of the Robert Koch Institute. Calendar week 26/2021. https://www.rki.de/EN/Content/infections/epidemiology/outbreaks/COVID-19/Situationsberichte_Tab.html

